# Year-long assessment of the immune response elicited by MVA-BN vaccine

**DOI:** 10.1101/2024.09.20.24313813

**Authors:** Giulia Matusali, Eleonora Cimini, Valentina Mazzotta, Francesca Colavita, Fabrizio Maggi, Andrea Antinori

## Abstract

**Background:** Modified-Vaccinia-Ankara Bavarian Nordic (MVA-BN) vaccine has been recommended to tackle the mpox epidemic 2022-2023 and its resurgence in 2024.

Although its effectiveness has been estimated to range between 36-86%, the persistence of protection is still unknown.

Aims of this study is to assess the immune response one year after vaccination with MVA-BN.

**Methods:** Observational prospective study at the National Institute for Infectious Diseases in Rome. All people at high risk for mpox infection who received MVA-BN as pre-exposure prophylaxis were enrolled. People previously primed with smallpox vaccination received a single-dose course of MVA-BN, while non-primed received a two-dose course. Blood samples were collected at the time of each dose and one, six and 12 months after vaccination. MPXV-specific IgG and neutralizing antibody (nAb) titers were assessed by immunofluorescence and plaque neutralization tests, respectively. Interferon-γ producing T-cell specific response to the MVA-BN vaccine was analyzed by ELISpot assay. Antibody titers at pre- and post-vaccination were compared using the Friedman tests. Mann Whitney test was used to compare antibody titers in PLWH vs PLWoH. Wilcoxon and Mann-Whitney non-parametric tests were used to compare T-cells specific response to the MVA-BN vaccine for intra and inter-group differences, respectively.

**Results:** Fifty high-risk people were included. All were men, with 94% self-reporting having sex with men. The median age was 50 years (IQR 45-57), and 21 (42%) were people living with HIV (PLWH), all on antiretroviral therapy, and 71% with a CD4 cell count higher than 500 μL. 25 (50%) have been primed with previous smallpox vaccination.

In non-primed people, anti-MPXV IgG titers significantly increased from T1 to T3 and, despite a slight reduction, were still higher than T1 up to T4 and then gradually decreased until T5, when 64% of sera were still reactive. MPXV-nAb titers peaked at T3 and then dropped, with 56% and 32% of sera reactive at T4 and T5, respectively. IFN-γ production by MVA-BN-specific T-cells progressively rose across time, peaked at T3, and remained significantly higher than the baseline after 6 and 12 months from vaccination. A single-dose course of MVA-BN vaccination in smallpox-primed participants elicited an early increase in IgG and nAb titers, which remained significantly higher than baseline after 6 and 12 months. MPXV-nAbs were detected in 80% and 72% of vaccinees at T4 and T5, respectively. A similar improvement and maintenance were observed for the MVA-BN-specific T-cell response. No evidence for a difference in both humoral and cellular responses was found between PLWH and PLWoH in our cohort.

**Conclusions:** One year after vaccination, our data showed the persistent detectability of low levels of nAb against MPXV in one-third of non-primed individuals. At the same time, humoral response was still detectable in most previously vaccinated participants. Concurrently, the MVA-BN-specific T-cell response was robust and persistent.

## Main text

Since May 2022, nearly 100,000 mpox cases have been reported.^1^ Currently, the rise of new cases in Africa, with the spread of the new MPXV Clade Ib together with the Clade IIb, led the World Health Organization to declare mpox as a public health emergency of international concern again.^2^ Modified-Vaccinia-Ankara Bavarian Nordic (MVA-BN) vaccine has been recommended to tackle the epidemic. According to epidemiological models, its effectiveness ranged between 36-86%.^3-6^ MVA-BN was reported to be safe and immunogenic early after vaccination^7^ and to elicit a strong cellular response, while a moderate humoral one. ^8,9^ Raising concerns about the possible spread of a new epidemic have posed the question of protection persistence after vaccination, but limited data are available on the duration of immunity^10^. Here, we describe the kinetics of humoral and cellular immune responses up to one year after vaccination.

Fifty high-risk people vaccinated at the Lazzaro Spallanzani Institute in Rome, Italy, during the 2022-2023 outbreak were included. All were men, with 94% self-reporting having sex with men. The median age was 50 years (IQR 45-57), and 21 (42%) were people living with HIV (PLWH), all on antiretroviral therapy, and 71% with a CD4 cell count higher than 500 μL. 25 (50%) have been primed with previous smallpox vaccination and received a single-dose course of immunization (Appendix 1). Blood samples were prospectively collected at the time of each dose administration (T1-T2) and then one (T3), six (T4), and twelve (T5) months from vaccination. Protocol and laboratory assays were previously described ^7^ (Appendix 2).

In non-primed people, anti-MPXV IgG titers significantly increased from T1 to T3 and, despite a slight reduction, were still higher than T1 up to T4 and then gradually decreased until T5, when 64% of sera were still reactive (Appendix 3). MPXV-nAb titers peaked at T3 and then dropped, with 56% and 32% of sera reactive at T4 and T5, respectively. IFN-γ production by MVA-BN-specific T-cells progressively rose across time, peaked at T3, and remained significantly higher than the baseline after 6 and 12 months from vaccination. (Figure 1).

**Figure 1:**
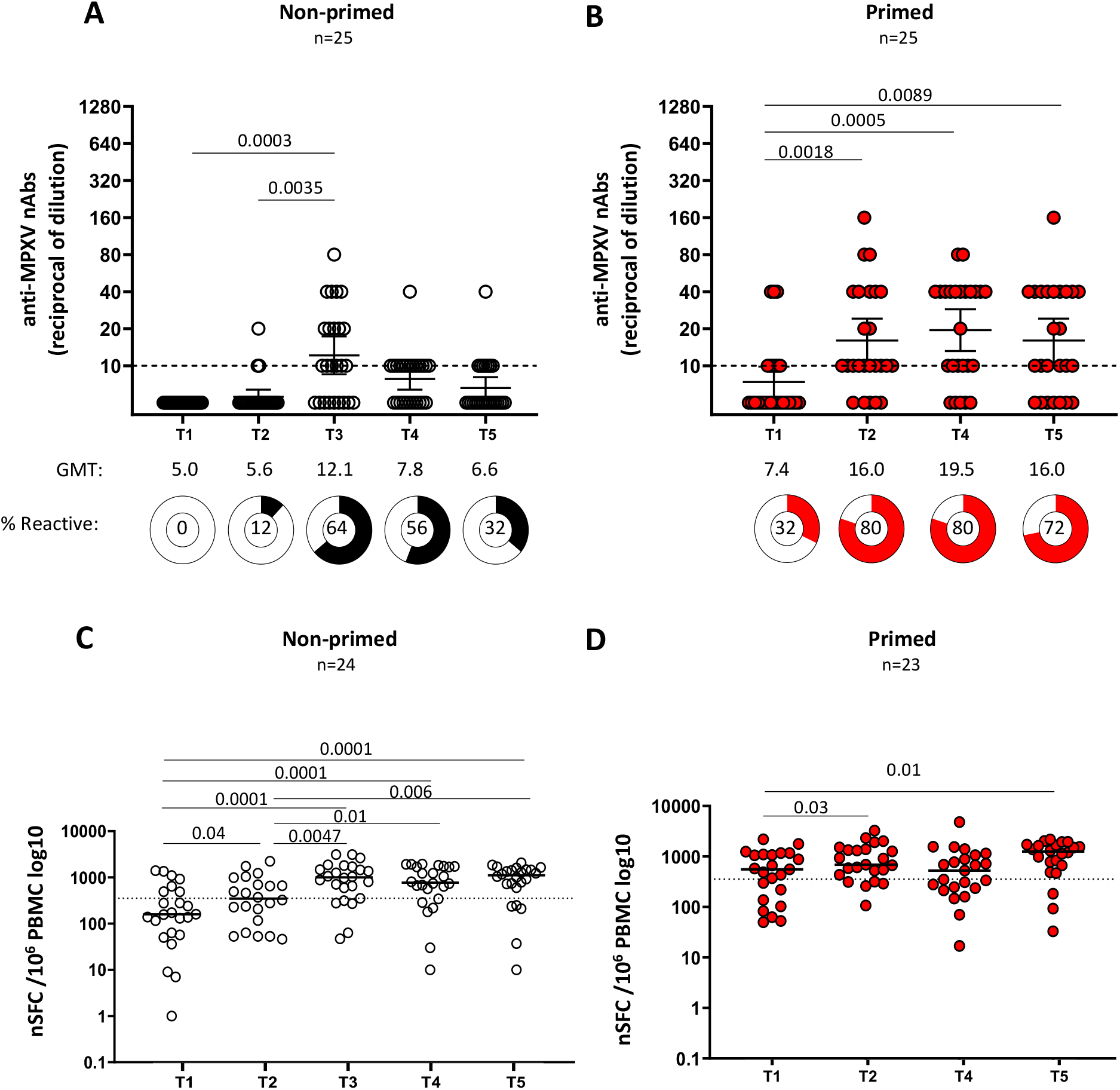
Kinetics of the neutralizing and cellular immune response in non-primed and primed MVA-BN vaccinated individuals. Panels A and B show the kinetics of anti-MPXV neutralizing antibody levels. The Dot line represents the limit of the assay detection, GMT= geometric mean titre. Panels C and D show the kinetics of T-cell-specific response after stimulation with MVA-BN. Black dots represent non-primed individuals; red dots represent primed individuals. Friedman and Wilcoxon’s tests were used for statistical comparisons.

Furthermore, a single-dose course of MVA-BN vaccination in smallpox-primed participants elicited an early increase in IgG and nAb titers, which remained significantly higher than baseline after 6 and 12 months (Figure 1 and Appendix 3). Notably, MPXV-nAbs were detected in 80% and 72% of vaccinees at T4 and T5, respectively. We observed a similar improvement and maintenance in the MVA-BN-specific T-cell response (Figure 1). No evidence for a difference in both humoral and cellular responses was found between PLWH and PLWoH in our cohort (Appendix 4). Limitations are stated in Appendix 5.

One year after vaccination, our data showed the persistent detectability of low levels of nAb against MPXV in one-third of non-primed individuals. At the same time, humoral response was still detectable in most previously vaccinated participants. Concurrently, the MVA-BN-specific T-cell response was robust and persistent. Whether this T-cell response can contribute to ensuring long-term protection against mpox, even in individuals with a significant reduction in the humoral response, remains to be demonstrated. This will be better clarified through epidemiological studies and longer follow-ups of vaccinated individuals. These data may be informative for public health recommendations on the need and time for booster doses after a primary course of MVA-BN^7^.

## Data Availability

All data produced in the present study are available upon reasonable request to the authors

## Appendix

**Table 1.**
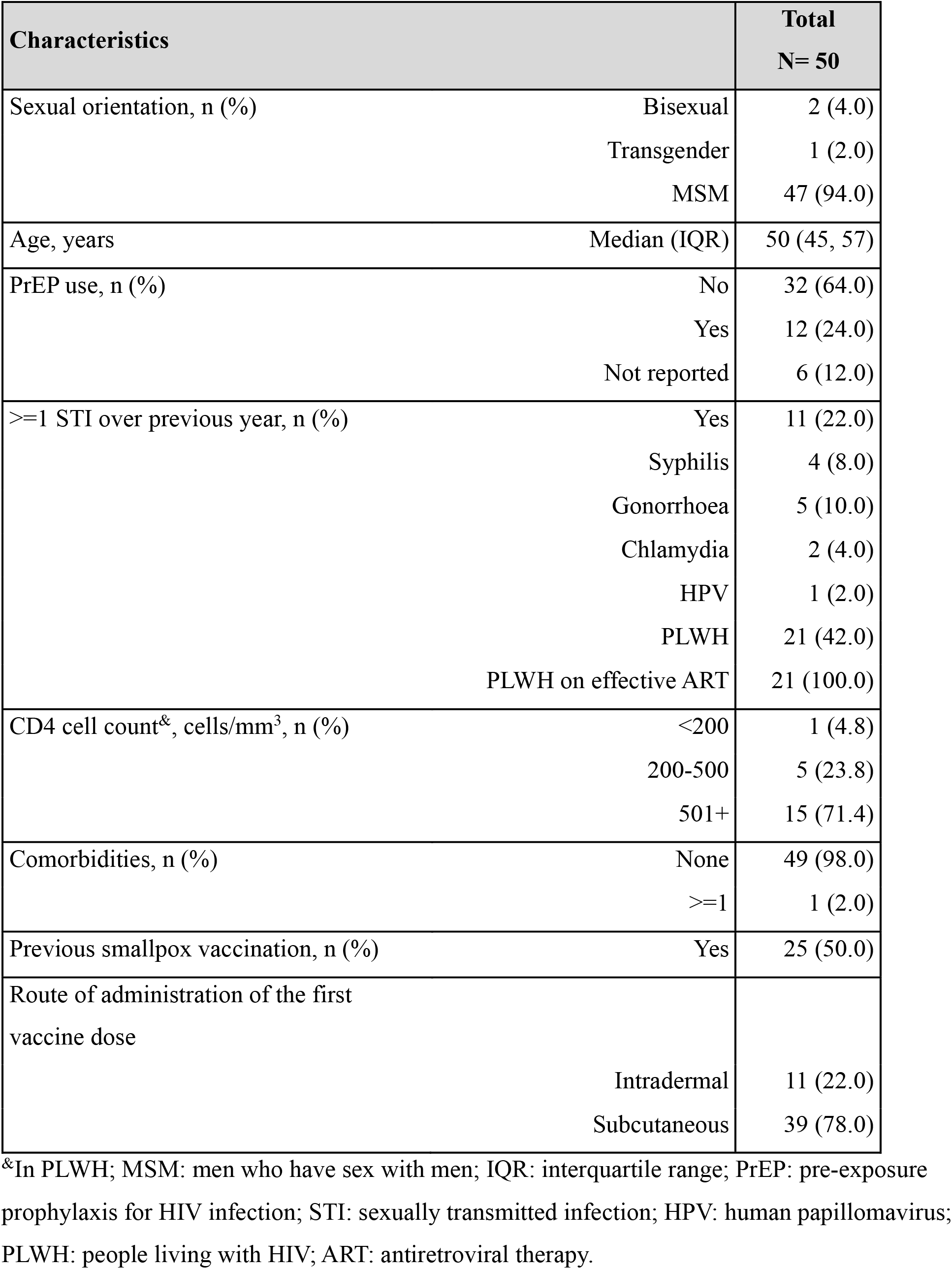
Characteristics of the study population.

## 2 Study protocol and Methods

### Study protocol

Mpox-Vac study (“*Studio prospettico osservazionale per monitorare aspetti relativi alla sicurezza, all’efficacia, all’immunogenicità e all’accettabilità della vaccinazione anti Monkeypox con vaccino MVA-BN (JYNNEOS) in persone ad alto rischio*”) was approved by the INMI Lazzaro Spallanzani Ethical Committee (approval number 41z, Register of Non-Covid Trials 2022). All subjects eligible for mpox vaccination according to the Italian Ministry of Health guidelines^1^ and who signed a written informed consent were enrolled in the study. At baseline (when receiving the first or single MVA-BN dose, T1), subjects were evaluated for demographic and behavioural characteristics linked to mpox exposure. Information regarding HIV status, CD4 count, HIV pre-exposure prophylaxis (PrEP) uptake and any history of previous STIs was collected. Other time points were scheduled at the second dose (T2) and then one, six and 12 months after the completion of the cycle (T3-T4-T5). For the vaccine-experienced individuals (primed) who received a single-dose schedule as a complete vaccination cycle, T2 was the time point corresponding to one month after vaccination completion. At each time point, blood samples were collected for the assessment of the immune response.

### Laboratory methods

Anti-MPXV IgG in serum were titered using an indirect immunofluorescence assay (IFA) based on in-house prepared slides with Vero E6 cells infected with an MPXV isolated from the skin lesion of a mpox patient hospitalized during the 2022 outbreak (GenBank: ON745215.1). The starting dilution for the testing of serum samples was 1:20. The secondary antibody was purchased from Euroimmun.

For measuring anti-MPXV nAb, serum samples were heat-inactivated at 56°C for 30 min and titrated in duplicate in 4 four-fold serial dilutions (starting dilution 1:10). Each serum dilution was added to 100 TCID50 MPXV isolate (GenBank: ON745215.1) and incubated at 37°C for 2 h. Subsequently, 96-well tissue culture plates with Vero E6 cell monolayers were infected with 120 μL/well of virus/serum mixtures and incubated at 37°C and 5% CO2. After 5 days, the supernatant was discarded, a staining-fixing solution (crystal violet containing 10% formaldehyde Diapath S.P.A. and Sigma-Aldrich) added for 30 min, removed, and washed with PBS 1X (Sigma-Aldrich). The number of plaques was counted using the Cytation 5 reader (Biotek) and when the average number of plaques was lower than half the number counted in the virus alone, serum sample was considered neutralizing. The highest serum dilution showing at least 50% of the plaques number reduction was indicated as the 50% neutralization titer (PRNT50%). Each test included serum control (1:10 dilution of each sample tested without virus), cell control (Vero E6 cells alone), and virus control (100 TCID50 MPXV in sextuplicate).

Peripheral blood mononuclear cells (PBMC) were isolated from enrolled subjects using Ficoll density gradient centrifugation (Pancoll human, PAN Biotech) and frozen in FBS (Fetal bovine serum, Euroclone, Italy) added of 10% DMSO (Merck Life sciences, Milan, Italy) at vapors of liquid nitrogen for the experimental design.

Interferon-γ producing T-cell specific response to the MVA-BN vaccine was analyzed by ELISpot assay. PBMC were thawed and suspended in complete medium [RPMI-1640 added of 10% fetal bovine serum, 1% L-glutamine, and 1% penicillin/streptomycin (Euroclone, Italy)]. Live PBMC were counted by Trypan blue exclusion and plated at 3×10^5^ cells/well in ELISpot plates (Human IFN-y ELISpot plus kit; Mabtech, Sweden). MVA-BN vaccine suspension (MOI 1) [JYNNEOS (Smallpox and Monkeypox Vaccine, Live, non-replicating)] added of αCD28/αCD49d (1 µg/ml, BD Biosciences) was used for 20h stimulation at 37 °C (5% CO_2_). After the incubation, the ELISpot assay was developed following manufacturer’s instructions. The spontaneous IFN-γ release was calculated in unstimulated culture (background) and a superantigen (SEB, 200nM, Sigma) was used as positive control. Results are expressed as spot-forming cells per 10^6^ PBMC (SFC/10^6^ PBMC).

### Statistical analysis

Antibody titers at different time points pre- and post-vaccination were compared using the Friedman tests. Mann Whitney test was used to compare antibody titers in PLWH vs PLWoH. Wilcoxon and Mann-Whitney non-parametric tests were used to compare T-cells specific response to the MVA-BN vaccine for intra and inter-group differences, respectively.

Analyses were performed using GraphPad Prism version 10 (GraphPad Software) for Windows statistical software; p < 0.05 was considered statistically significant.

**Figure 1:**
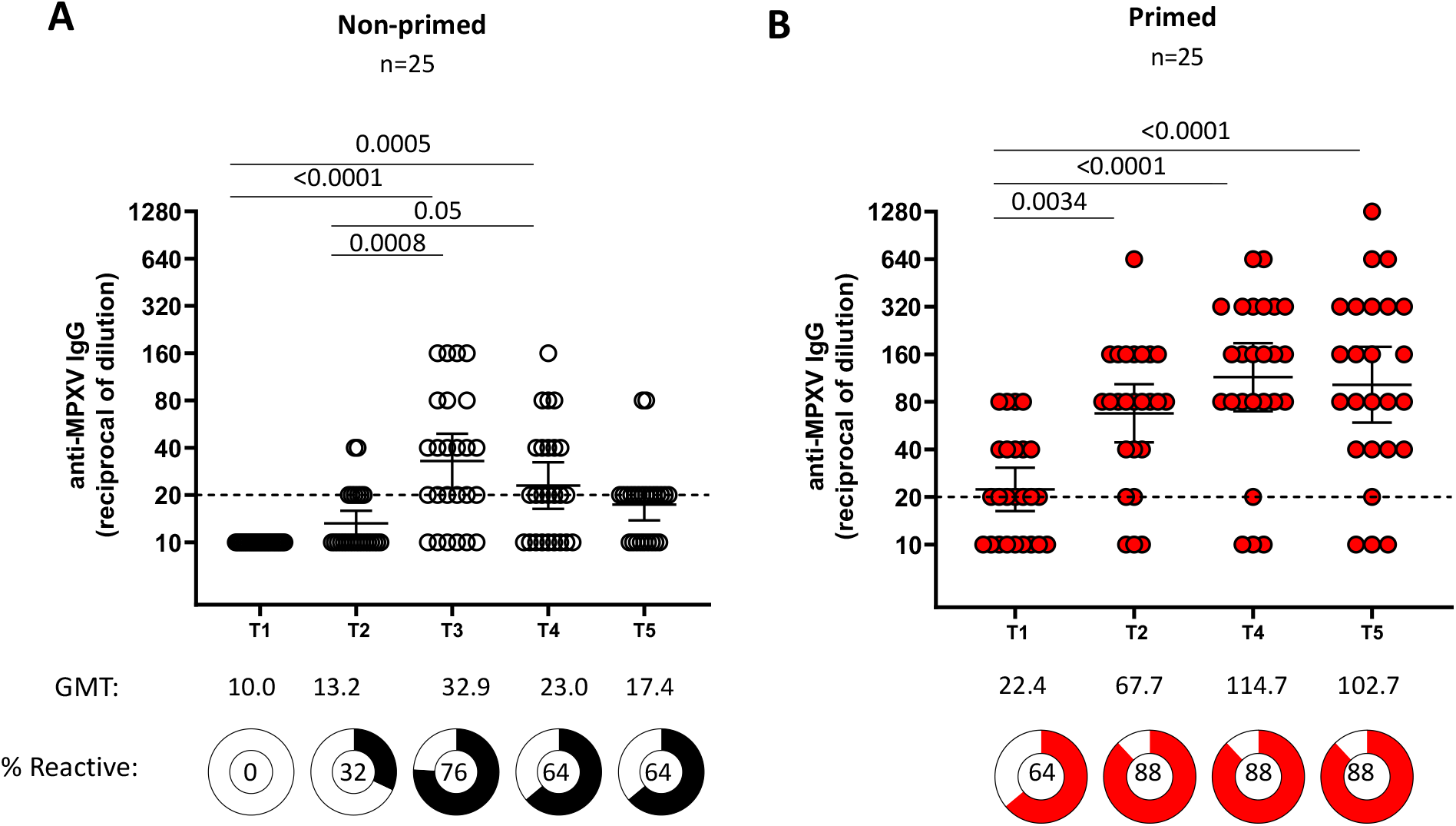
Kinetics of IgG MPXV-specific antibodies (IgG) in non-primed (Panel A) and primed (Panel B) individuals. Titers of MPXV-specific IgG were measured by immunofluorescence (1:20) starting dilution. The Dot line represents the limit of the detection of the assay. Titers are expressed as Geometric Mean Titres (GMT) of the reciprocal serum dilution (log2 scale). Error bars refer to the 95% confidence interval of GMT.

**Figure 2:**
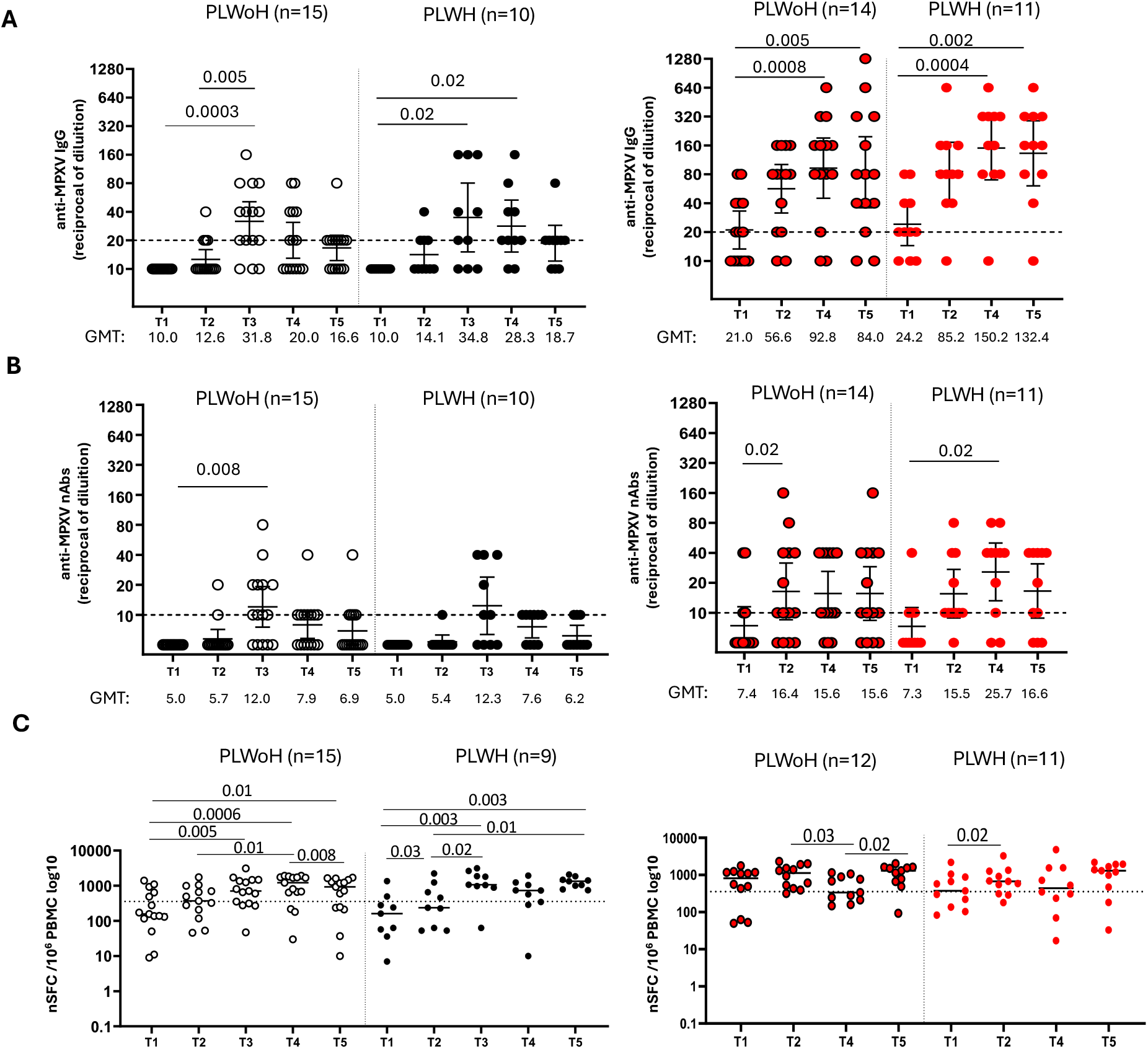
Humoral and cellular response according to HIV status. Kinetics of MPXV-specific IgG (Panel A), neutralizing antibodies (Panel B) and T-cell specific (Panel C) after MVA-BN vaccination, according to HIV status. Mann-Whitney test was used to compare people living with (PLWH) and without HIV infection (PLWoH). Black dots represent non-primed individuals; red dots represent primed individuals.

## 5 Limitations

The observational nature of the study could have introduced bias in the selection of participants for the study. We acknowledge the limited sample size, which might have limited the statistical power for some comparisons (e.g. PLWH vs. PLWoH). However, blood samples were prospectively collected, and the time points were respected.

## 7 Mpox Vaccine Lazio Study Group

C Aguglia, A Antinori, E Anzalone, A Barca, M Camici, F Cannone, P Caputi, R Casetti, L Caterini, C Cimaglia, E Cimini, F Colavita, L Coppola, R Corso, F Cristofanelli, S Cruciani, N De Marco, G Del Duca, G D’Ettorre, S Di Bari, S Di Giambenedetto, P Faccendini, F Faraglia, D Farinacci, A Faticoni, C Fontana, M Fusto, R Gagliardini, P Gallí, S Gebremeskel, G Giannico, S Gili, E Girardi, G Grassi, MR Iannella, A Junea, D Kontogiannis, A Lamonaca, S Lanini, A Latini, D Lapa, M Lichtner, MG Loira, F Maggi, A Marani, M Marchili, R Marocco, A Masone, C Mastroianni, I Mastrorosa, G Matusali, V Mazzotta, S Meschi, S Minicucci, A Mondi, V Mondillo, A Nappo, G Natalini, E Nicastri, S Notari, A Oliva, A Parisi, J Paulicelli, C Pinnetti, P Piselli, MM Plazzi, A Possi, G Preziosi, R Preziosi, G Prota, M Ridolfi, S Rosati,, A Russo, L Sarmati, P Scanzano, L Scorzolini, C Stingone, E Tamburrini, E Tartaglia, V Tomassi, F Vaia, A Vergori, M Vescovo, S Vita, J Volpi, P Zuccalà.

## REFERENCES

1. Centers and Disease Control and Prevention: 2022 -2023Monkeypox Outbreak Global Map. 2023, Data as of August 6th 2024 at 5:30 pm EDT, accessed on August 9th, 2024.

2. https://www.who.int/news/item/14-08-2024-who-director-general-declares-mpox-outbreak-a-public-health-emergency-of-international-concern

3. Deputy NP, Deckert J, Chard AN, et al. Vaccine Effectiveness of JYNNEOS against Mpox Disease in the United States. N Engl J Med. 2023;388(26):2434–2443.

4. Payne AB, Ray LC, Cole MM, et al. Reduced Risk for Mpox After Receipt of 1 or 2 Doses of JYNNEOS Vaccine Compared with Risk Among Unvaccinated Persons - 43 U.S. Jurisdictions, July 31-October 1, 2022. MMWR Morb Mortal Wkly Rep. 2022;71(49):1560–1564.

5. Bertran M, Andrews N, Davison C, et al. Effectiveness of one dose of MVA–BN smallpox vaccine against mpox in England using the case-coverage method: an observational study. Lancet Infect Dis. 2023;Mar 13.

6. Wolff Sagy Y, Zucker R, Hammerman A, et al. Real-world effectiveness of a single dose of mpox vaccine in males. Nat Med. 2023; 29(3):748–752.

7. Mazzotta V, Lepri AC, Matusali G, et al. Immunogenicity and reactogenicity of modified vaccinia Ankara pre-exposure vaccination against mpox according to previous smallpox vaccine exposure and HIV infection: prospective cohort study. EClinicalMedicine. 2024 Jan 12;68:102420.

8. Cohn H, Bloom N, Cai GY, et al. Mpox vaccine and infection-driven human immune signatures: an immunological analysis of an observational study. Lancet Infect Dis. 2023;S1473-3099(23)00352-3.

9. Zaeck LM, Lamers MM, Verstrepen BE, et al. Low levels of monkeypox virus-neutralizing antibodies after MVA-BN vaccination in healthy individuals. Nat Med. 2023; 29(1): 270–278.

10. UK Health Security Agency. Recommendations for the use of pre- and post-exposure vaccination during a monkeypox incident. Updated 26 August 2022 v12; https://assets.publishing.service.gov.uk/media/6308acef8fa8f55363756beb/recommendations-for-pre-and-post-exposure-vaccination-during-a-monkeypox-incident-26-august-2022.pdf

## References in Appendix

1 Ministero della Salute. Circolare del Ministero della Salute n. 35365 - Indicazioni ad interim sulla strategia vaccinale contro il vaiolo delle scimmie (MPX). (2022).

